# Getting to the cause of Chronic Kidney Disease of unknown cause (CKDu): Research protocol and baseline results

**DOI:** 10.64898/2026.04.30.26352115

**Authors:** Marvin Gonzalez-Quiroz, Aurora Aragón, Prabhdeep Kaur, Sharan Murali, Manikandanesan Sakthivel, Thilanga Ruwanpathirana, Pubudu Chulasiri, Nalika Gunawardena, Charlotte E Rutter, Ben Caplin, Neil Pearce

**Author notes:** <u>Address for correspondence:</u> Professor Neil Pearce, Department of Medical Statistics, London School of Hygiene and Tropical Medicine, London, United Kingdom.

## Abstract

There is an epidemic of primarily tubular-interstitial chronic kidney disease (CKD) clustering in agricultural communities in low-and-middle income countries (LMICs). Although it is currently unclear whether there is a common underlying cause, these conditions have been collectively termed CKD of unknown cause (CKDu). CKDu is estimated to have led to the premature deaths of tens to hundreds of thousands of young adults in LMICs over the last two decades. Thus, there is an urgent need to understand the aetiology and pathophysiology of these conditions and to develop preventive interventions.

We have now established that CKDu exists in Central America (Nicaragua) and South Asia (India, Sri Lanka), but not in some other tropical countries. It is not clear yet whether the epidemics in Central America and South Asia have common causes or different causes, which is why it is important to conduct research using the same protocols and methods in these different regions.

We have therefore established prospective studies in affected communities in Nicaragua, South India, and Sri Lanka to investigate the causes of the epidemics of CKDu, and factors which affect prognosis. The underlying hypothesis is that CKDu is caused by unknown factors to which the populations have become exposed, due to changes in agricultural practice or other environmental changes (e.g. water supply), over recent decades.

The objectives of the collaboration are to investigate the environmental causes of renal decline in these high-risk populations, using standardised instruments capturing occupational and environmental exposures. We will address four proposed causes of CKDu: (i) metals and metaloids; (ii) agrochemicals; (iii) infections by organisms that affect the kidney; and (iv) heat/dehydration.

## Introduction

There is a high prevalence of primarily tubular-interstitial chronic kidney disease (CKD) in agricultural communities in low-and-middle income countries (LMICs)(1, 2). These ‘epidemics’ involve CKD in the absence of the established risk factors of diabetes, primary glomerular nephritis and structural abnormality. This form of CKD has therefore been termed CKD of unknown cause (CKDu). CKDu is clinically silent, usually diagnosed late, and no treatment. Renal replacement therapy (RRT) is limited in many LMICs(3), especially in the affected areas, where progression to end stage renal disease (ESRD) often leads to premature death. In the last few decades, it has been a leading cause of death among adult 18-38 years old in several of the affected countries(4). Variation in access to RRT, and variation in hospital coding variability, means that routine health data are not accurate, or representative. Therefore, valid prevalence estimates can only be obtained by representative population surveys(5). In a series of investigations, we have shown that a high burden of CKDu exists in Central America(6), India and Sri Lanka(7). It may also be occurring in other tropical/subtropical regions where comparable data are not currently available(2, 8), but to date no ‘epidemic areas’ have been identified in population-based surveys outside of Central America or South Asia (9).

A large number of possible causes have been suggested for CKDu. <u>Heat/dehydration</u>, <u>infections</u> and <u>pesticides</u> are the main hypotheses in Central America, whereas in India and Sri Lanka the emphasis has been on <u>heavy metals in water</u> and/or pesticides.(7, 10) Other possible causes include misuse of non-steroidal anti-inflammatories (NSAIDs), population genetics and alcohol(4). Biopsy studies have been helpful in excluding known causes of kidney failure, but have provided few clues to the aetiology(11). The most consistent finding is that CKDu is more common in agricultural communities(12). In Central America, CKDu occurs frequently in sugar cane workers, but also in other agricultural workers, fishermen, miners, and construction workers including brickmaking(13-15); it also occurs in some women in the same areas, most of whom have not worked in agriculture(16). We conducted a systematic review of studies in Central America: the only consistent risk factors were male sex, family history of CKD, high water intake, and low altitude(4). Heat stress is strongly associated with established disease, and with disease progression, but is unlikely to be the primary cause(17). Other hypotheses, including pesticide exposure, alcohol, and NSAIDS, have little or no support to date. However, there have been few studies involving biomarkers of exposure or pathophysiology.

### The need for multi-centre prospective studies

Although population-based surveys provide useful high-level data on the presence or absence of endemic CKDu, this design has shortcomings when investigating the aetiology of CKDu.

Firstly, identifying early disease is key as there are a wide range of common factors in low-income settings, e.g. episodes of dehydration and widespread use of NSAIDs, which may exacerbate GFR decline once CKD (of any cause) is established, but may not be the primary causes. This, alongside issues of reverse causation and recall bias, means that cross-sectional and case-control approaches have not been useful in uncovering the primary cause of disease. Furthermore, one-off eGFR measures show little utility in identifying early disease, because of: (i) substantial non-renal variation in eGFR between healthy individuals and; (ii) short-term reversible changes in renal function within individuals. Alongside the absence of urinary abnormalities this means within-individual decline in eGFR in the normal, or near normal range, is currently the only way to detect early cases.

Secondly, biopsies from all three regions have failed to uncover the cause of disease, but show substantial similarities, that is, interstitial fibrosis, tubular atrophy, glomerular ischaemia and sclerosis(18), findings which are consistent with kidneys subject to wide range of tubular or vascular insults, but tell us little about the initiating exposure. For example, a recent report(19) of tubular cytoplasmic inclusions, proposed evidence for a calcineurin-inhibitor type toxin, but these findings have been found to be non-specific(20, 21), perhaps because biopsies are performed at later stages of disease and the initiating factor is long gone. Alternatively, reports of acute inflammation in biopsies during febrile illness in otherwise healthy patients from high-risk areas in both Sri Lanka(22) and Nicaragua(23) might support an infective cause, but associations with such clinical syndromes with CKDu can only be explored with longitudinal studies with adequate population controls. Furthermore, renal biopsy is logistically challenging in the study regions and our pragmatic clinical criteria for disease (age, declining eGFR, absence of diabetes, hypertension and heavy proteinuria), have high positive predictive value, i.e. no alternative histological diagnoses have been identified in patients meeting those criteria in biopsy studies in the endemic regions(11). In turn suggesting biopsy confirmation of cases is unnecessary for research purposes in epidemic areas and would add complexity without substantial benefit.

### Prospective studies in Nicaragua, India and Sri Lanka

Therefore, we have now established three prospective studies of within-person change in eGFR over time in areas where we have established that CKDu is endemic. The three cohorts are based on two high prevalence areas in Anuradhapura, Sri Lanka, the high prevalence area in Andhra Pradesh, South India, and the high prevalence areas in Pacific Coast Nicaragua; Population-based surveys in these settings have reported a prevalence of low eGFR of 5-10% in working age adults without diabetes and hypertension. The characteristics of the cohort study participants are given in Table 1.

**Table 1:** Demographic characteristics of baseline populations by sex.

|  | Nicaragua (n=1001) |  | India (n= 206) |  | Sri Lanka (n=729) |  |
| --- | --- | --- | --- | --- | --- | --- |
| Variable | Males (n=598) | Females (n=403) | India males (n=84) | India females (n=122) | Sri Lanka males (n=407) | Sri Lanka females (n=322) |
| Age (yrs), mean ± SD | 22.73± 3.72 | 23.04± 3.56 | 38.58 ± 11.56 | 38.49 ± 11.27 | 35.82 ± 10.21 | 36.06 ± 9.38 |
| Age-range (years) | 18 – 30 | 18 - 30 | 18–60 | 18–60 | 17 – 56 | 18 – 59 |
| Years of school, mean ± SD | | | 5.19 ± 5.32 | 2.66 ± 4.23 | | |
| Passed school exams at age 16, n (%) |  |  |  |  | 229 (56.3) | 184 (57.3)<br>(missing=1) |
| Number of family members, median (IQR) | 4.31 + 1.9 | 3.98 + 1.61 | 2 (2, 3) | 2 (1, 2) |  |  |
| Body mass index (BMI), median (IQR) | 22.29 (20.69 – 24.40) | 25.53 (22.38 – 29.84) | 22.30 (19.90, 24.77) | 21.30 (18.60, 24.25) | 21.63 (18.56, 24.71)<br>(missing=1) | 22.84 (19.48, 26.04) |
| Systolic blood pressure (mmHg), median (IQR) | 121 (112 – 128) | 111 (105 – 119) | 127.67 (115.83,140.25) | 119.17 (112.00, 130.33) | 115.5 (107.5, 124) | 112.3 (104, 121)<br>(missing=2) |
| Diastolic blood pressure (mmHg), median (IQR) | 71 (65 – 77) | 70 (66 – 76) | 84.50 (77.67, 93.33) | 80.67 (75.50, 86.67) | 70.5 (65, 77.5) | 71 (65.5, 77.5)<br>(missing=2) |
| Heart rate, median (IQR) | 69 (63 – 77) | 80 (73 – 88) | 78.17 (71.33, 86.83) | 77.67 (71.92, 84.33) |  |  |
| eGFR ML/min/1.73m<sup>2</sup>, median (IQR) | 123.38 (107.11 – 131.16) | 123.57 (105.73 – 129.80) | 102.23 (85.26, 117.27) | 108.57 (96.39, 119.31) | **104.43 (89.63, 116.29) | **97.82 (82.81, 110.12) |
| Ever smoked, n (%) | 272 (45.4) | 3 (0.7) | 29 (34.5%) | 0 (0.0%) | 237 (58.2) | 2 (0.6) |
| Ever drank alcohol, n (%) | 325 (54.3) | 32 (7.9) | 35 (41.7%) | 0 (0.0%) | 286 (71.9)<br>(missing=9) | 5 (1.7)<br>(missing=19) |
| NSAIDS, n (%) | 525 (87.7) | 377 (93.5) | 0 (0.0%) | 6 (4.9%) |  |  |
| Urinary tract infection, n (%) | 94 (15.7) | 111 (27.5) | 1 (1.2%) | 4 (3.3%) | *8 (2.0) | *10 (3.1) |
| Nephrotoxic antibiotics, n (%) | 40 (6.6) | 40 (9.9) | 0 (0.0%) | 6 (4.9%) |  |  |

\*ever had chronic urinary tract infection \*\*calculated using CKD-EPI 2021 for creatinine, some samples have since been reanalysed for longitudinal analysis.

#### Nicaragua

We established a prospective study in 11 rural communities in North-western Nicaragua. The study protocol has since been used more widely(24). After exclusions, Phase I (2014), recruited all eligible men (n=263) and a random sample of women (n=87), aged 18-30, without self-reported CKD, diabetes or hypertension(25). The baseline response rate was 97%(25), and retention among male participants during the first two years of follow-up was 92%(16). Phase II, conducted in 2018, enrolled an additional 213 men and 207 women. In 2021, during phase III, 224 new participants were recruited (114 men and 111 women) applying the same inclusion criteria as in Phase I, but targeting a 1:1 male-to-female ratio. Overall, retention across study phases remained high (85.9%).

#### Sri Lanka

We established a similar prospective study in Sri Lanka, following the same core protocol and collecting comparable risk factor information) to the Nicaragua study(24, 25), with funding from the World Health Organization, Country Office, Sri Lanka. This community-based prospective study included annual follow-up for at least four years. The study setting was identified based on the DEGREE surveys conducted in five areas of Anuradhapura district in 2017(26). The two areas with the lowest mean eGFR (Puhudivula and Lolugaswewa) were selected. The study population comprised healthy residents aged 20-60 years without evident kidney impairment, verified by serum creatinine measurement using IDMS (isotope dilution mass spectrometry) standards. Recruitment was in October 2018: 729 of 750 invited individuals participated (97%). The first annual follow-up, conducted in 2019 was completed by 709 (97.2%): the second, in 2021 by 703 (96.4%): and the third in 2023 by 716 (98.2%).

#### India

We have also established a similar prospective study in India, following the same core protocol (25). Villages with a high prevalence of chronic kidney disease were identified based on reports from local clinicians and dialysis data from the state-funded insurance programme in Prakasam district, Andhra Pradesh. Between December 2021 and August 2024, all households and individuals in the 18-60 years age group were enumerated in eight villages. The baseline survey included over 1637 residents between the ages of 18-60 years. We collected data on risk factors, previous disease history, anthropometric measurements and blood pressure measurements. Based on the survey, we identified two geographically contiguous villages with a CKDu prevalence of more than 5%. We have identified 206 individuals without obvious kidney function impairment, verified by estimation of serum creatinine using IDMS standards, for the cohort study. At the time of writing, three follow-up rounds had been completed, with a response rate of approximately 70% across rounds. Additional adjacent villages are being surveyed to identify further high-prevalence communities, with the aim of expanding the Indian cohort to at least 1,000 participants without evident kidney impairment.

We are now standardizing, integrating and developing further these three cohort studies to establish a comprehensive programme of work, with a common protocol to identify the cause(s) of CKDu in these areas.

### Study protocol

The underlying hypothesis is that CKDu is caused by currently unknown environmental factors in each endemic region. While the key risk factors may be different in each region, several exacerbating factors such as strenuous work in hot environments and limited access to healthcare are also likely to contribute, and may account for the similarities in the populations affected.

The objectives of this research are therefore to:

1. Investigate the environmental causes of kidney function decline in these high-risk populations (Nicaragua, India, Sri Lanka), using standardised instruments capturing occupational and environmental exposures specifically addressing four proposed causes of disease:
  a. Metals and metalloids (using direct measurement in biological samples)
  b. Agrochemicals (using direct measurement in biological samples)
  c. Infection by nephrotropic pathogens (using serological and metagenomic analyses)
  d. Heat/dehydration (estimating exposure in individuals using a heat-exposure-matrix)
2. Identify early biomarkers of disease and genetic risk loci to provide insight into disease biology and potential mechanisms underlying rapid decline in kidney function
3. Provide a bioresource of urine and blood samples from three robustly characterised prospective studies to allow the exploration of other causal hypotheses going forward.

#### Research plan

The collaboration involves three prospective studies. Comparable questionnaire information and biomarkers will be collected and tested, and standardized laboratory analyses will be conducted.

#### Workstream 1: Expansion and standardization of existing studies

##### Study Design

In each centre (Nicaragua, India, Sri Lanka): a prospective study of healthy men and women followed for at least four years.

###### Inclusion criteria

Participants without a pre-existing clinical diagnosis or history of CKD, diabetes or hypertension were invited to participate in the study.

##### Study Visits

All data are collected, where possible, using Open Data Kit (ODK) and tablet devices. Height (first visit only), weight and blood pressure (average of 3 measures following 5 minutes seated rest using a digital sphygmomanometer) are measured at each study visit using digital calibrated devices.

##### Questionnaire

An exposure questionnaire is administered by trained interviewers prior to each study visit. All participants will answer baseline questions on demographics, social and medical history, occupational, infectious (e.g. self-reported disease presentation) and environmental exposures including exposure to potential phytotoxins (e.g. aristolochia) using images of the implicated species, hydration patterns, medication use, low birthweight and prematurity.

##### Heat stress

Heat stress is perhaps the most difficult exposure to measure as there are currently no good biomarkers of chronic heat stress. We will use: (i) standard questionnaires on symptoms of heat stress(16, 25); (ii) an additional detailed questionnaire on time spent in various locations (particularly home and work) and activities (including work practices); and (iii) climate data (already available with a high resolution from the climate reanalysis ERA-5, which provides global and free data climate variables on a 30 km resolution grid from 1979 to present) on local temperature, humidity, etc, during each month of the year. This information will be used for a heat-exposure-matrix (similar to a job-exposure-matrix, an approach which is commonly used in occupational studies(27)), to estimate each participant’s chronic exposure to heat and heat stress.

##### Biospecimens

Urine and blood samples are collected at each study visit. Urinalysis is performed on site using dipstick tests. Blood samples were processed at the point of collection: serum was separated using mobile centrifuges and whole blood samples were stored in an icebox at 4°C. Samples were transported to the study centre, where biospecimens were stored at -80C (except the Nicaraguan samples which were stored at -20°C prior to transfer to the UK within two weeks).

##### Outcome Assessment

Serum creatinine will be measured locally after each study visit to provide participants with timely information on kidney function. Along with samples from the final study visit a subset of samples from baseline and each follow-up visit will then be re-quantified in a single batch using an IDMS traceable reference method. The primary outcome is the eGFR trajectory over time, with the estimates calculated using CKD-EPI_SCr_ equation, (to minimize bias at well-preserved GFR)(28). Each laboratory is taking part in an external quality assurance scheme and study-specific eQA samples will be shared between the sites.

#### Workstream 2: Biomarkers of exposure

Testing will be conducted at HSE Science and Research Centre for urine samples collected in Nicaragua and Sri Lanka, and locally for the Indian samples. The Biological Monitoring team at HSE Science and Research Centre will make multi-sample external quality assurance samples available to external labs to ensure comparability.

##### Metals

Metals and metalloids in urine samples will be analysed using Inductively-Coupled Plasma – MS, including, aluminium, arsenic, cadmium, cobalt, copper, chromium, manganese, lead, selenium, silicon, silica, strontium and mercury.

##### Agrochemicals

A range of unmodified chemicals (e.g. N-(phosphonomethyl) glycine; glyphosate) and metabolites (e.g. of pyrethroid insecticides, 3-phenoxybenzoic acid, of pyrimetanil, 3-hydroxy-pyrimetanil) will be analysed in urine samples. Samples will either: (i) be de-conjugated using β-glucuronidase/arylsulphatase and prepared using solid phase extraction; (ii) be hydrolysed using alkaline conditions; or (iii) will use acidified and then quantified liquid chromatography-triple quadrupole linear ion trap MS. *Mycotoxins:* citrinin and ochratoxin will be quantified using established methodology(29).

##### Infectious serology

We will test serum samples collected at baseline in a subset of nested cases and controls by enzyme-linked immunosorbent assay (ELISA, e.g. Euroimmun) to measure immunoglobulin G (IgG) antibodies against locally prevalent hantaviruses and *Leptospira spp* at serial study visits. These pathogens were selected as they are locally endemic (for many years), known to cause acute kidney injury (AKI), and exposure is common in tropical agricultural settings. IgG seroconversion will be used to identify incident infections.

#### Workstream 4: Molecular biological disease insight

##### Genotyping

In common with other NCDs, the risk of developing CKDu will likely involve gene-environment interactions. Larger GWAS studies have been conducted of CKD(30), but these have been in Western populations where CKDu is not endemic or due to a single cause. We will aim to identify novel genetic loci which increase the risk of eGFR decline in these populations, as well as assessing loci that have previously been found to be important for CKD in general. Finding common genetic risk loci across sites would provide strong evidence for a unified aetiopathology of disease. Following DNA-extraction, genome wide variation will be captured using array technology (Ilumina Global Screening Array-24) and standard quality control applied. Given the close-knit communities from which the participants originate, a mixed-linear model association analysis(31) will be used to handle cryptic-relatedness/population stratification with change in eGFR over time (or case-control status as per the GMM below) as the outcome variable. Standard approaches will be used to assess systematic bias and to correct for multiple testing. We will also explore imputation methods to increase the power of this work.

##### Proteomics

Urinary proteins can provide insight into the functional segment of the nephron affected and mechanisms underlying kidney disease (32). Also, a urinary biomarker for CKDu is urgently required. Although the uACR is highly predictive of renal decline in most forms of CKD, albuminuria is typically completely absent until the late stages of CKDu (consistent with the tubular pathology observed on biopsy)(16). We have demonstrated that three eGFR measurements over 1-year are needed to reliably demonstrate early disease in the young population at-risk in Central America(33), hence absence of a one-off biomarker hampers aetiological research, clinical diagnosis and public health efforts. Therefore, we will perform discovery proteomics focusing on urine samples.

#### Workstream 5: Putting it all together

Once workstreams 1 to 4 are completed, we will conduct integrated analyses for each Centre, and pooled analyses across Centres. In each study, the integrative data will involve: (i) determinants of case status based on changes in eGFR using a range of statistical methods and (iii) nested case-control studies to identify risk factors and predictors case status. We will then conduct pooled analyses across the three studies, and where this is considered appropriate (i.e. if there are similar risk factors with similar effect sizes), we will produce pooled effect estimates using the approach described above. Each of the four main groups of potential causes (agrochemicals, metals, infections, heat stress), will be assessed together (with each exposure being controlled for the others). Some analyses will use causal modelling approaches, whereas others (e.g. biomarkers predictive of case status) will use predictive modelling(34).

##### Analysis of the full study data

In each study centre, we will initially analyze determinants of case status using the information (questionnaires, etc) which is available on all study members. For example, we will examine the association of any exposure with change in eGFR slope, estimated by the addition of variables (time-dependent where appropriate) to a baseline multi-level linear model or alternative statistical approaches. Alternative statistical methods will be used to identify disease onset.

##### Nested case-control analyses

Logistic regression models will be used to compare those with and without evidence of change in kidney function, adjusting for potential confounders using standard methods for modelling strategy, including checking for collinearity(34).

## Discussion

This protocol responds to a key problem that has limited progress in understanding CKDu, namely the absence of harmonized, longitudinal, multi-regional investigations capable of disentangling temporality, causality and context-specific exposures. Integrating prospective community-based cohort designs in these two endemic regions in Central America and South Asia will enable us, for the first time, to determine whether the condition has similar or different characteristics in the known endemic regions. It will also address long-standing methodological gaps that have hindered causal inference in CKDu research. Prior work has largely used cross-sectional surveys and retrospective assessments, which are inherently vulnerable to reverse causation, exposure misclassification, and others, thereby limiting their capacity to identify primary aetiological drivers.

The use of standardised protocols across geographically distinct countries enables direct comparison of exposure-outcome relationships, addressing a critical gap in the literature. To date, heterogeneity in study design, exposure assessment, and case definitions has precluded meaningful synthesis across regions. By harmonizing data collection and centralizing laboratory analyses, the laboratory methods allow for the identification of regionally specific risk factors while also testing the hypothesis of a shared aetiopathology.

A key strength of this protocol relies on its emphasis on within-individual trajectories of kidney function. Change in estimated glomerular filtration rate over time provides a more sensitive and specific indicator of early disease than single measurements, particularly in populations where baseline renal function is normal and albuminuria is typically absent until late stages. This aligns with the evidence that CKDu is characterized by a predominantly tubular-interstitial disease with minimal proteinuria, distinguishing it from traditional CKD pathways. By capturing longitudinal changes, this study is well positioned to differentiate initiating exposures from factors that merely accelerate progression, which has been difficult to establish in previous studies.

Despite these strengths, several challenges must be taken into consideration. In particular, loss to follow-up, particularly in economically vulnerable populations, could introduce selection bias, although the high retention rates reported in existing cohorts are reassuring. In addition, we did not collect longitudinal environmental samples (e.g., soil, dust and water), with the exception of Nicaragua, where water samples were collected during both dry and rainy seasons in the first year of the cohort (2014-2015). This limitation restricts our ability to compare toxicant levels between biological and environmental samples.

In conclusion, this protocol represents a methodologically rigorous and conceptually comprehensive approach to addressing one of the most pressing unresolved questions in nephro-epidemiology. By combining longitudinal data, standardised exposure assessment and molecular analyses across multiple endemic regions, it offers a unique opportunity to untangle the cause(s) of CKDu.

## Data Availability

All data produced in the present work are contained in the manuscript

## Acknowledgements

The authors thank the study participants and leaders from the study communities as well as the fieldwork teams in Nicaragua, India and Sri Lanka, without whom, none of this work would be possible. Nicaraguan investigators also acknowledge the contributions of the Research Centre on Health, Work and Environment (CISTA) for leading and hosting the project, FNE International for practical support, and members of La Isla Network for their input at the initial stages of this work. The Indian Co-investigators also acknowledge the valuable support and collaboration extended by the Noncommunicable Disease Cell at Prakasam District, Andhra Pradesh. We are especially grateful to the staff at Kanigiri Community Health Centre (CHC), including the dialysis unit staff, and the team at Primary Health Centre (PHC) Machavaram—comprising the Medical Officer, ASHA workers, ANMs, and other field staff—for their dedicated involvement in field operations. We sincerely thank Dr. Girish Chethrapilly Purushothaman Kumar, Scientist F and Head of the Laboratory Division, ICMR National Institute of Epidemiology, Chennai, for granting necessary permissions and providing institutional support for sample handling and storage. The authors also wish to thank Donna Davoren for technical and administrative assistance.

## Appendix 1: Power calculations

| Exposure groups | Risk factors (examples) | Likely prevalence | Likely effect size (continuous trait or nested case-control outcome)<sup>#</sup> | Power* (3 centres) | Power* (1centre) |
| --- | --- | --- | --- | --- | --- |
| Questionnaire (all participants) | Pesticides (occupational exposure) | 10%<br>20% | Additional eGFR decline of 5mL/min/year | >99%<br>>99% | >99%<br>>99% |
| Metals and metalloids (urine) (50 cases and 50 controls per centre) | Arsenic, aluminium, cadmium, silica, etc >75% percentile of exposure* | 25% | OR≥2.0<br>OR≥4.0 | 80%<br>>99% | 35%<br>92% |
| Agrochemicals (urine) (50 cases and 50 controls per centre) | Glyphosate, 2,4-D, MCPA, etc > 75% percentile of exposure | 25% | OR≥2.0<br>OR≥4.0 | 80%<br>>99% | 36%<br>92% |
| Infection by nephrotropic organisms (100 cases and 200 controls per centre) | Hantavirus, *leptospira spp.* | 12% | OR≥3.0<br>OR≥4.0 | 94%<br>>99% | 71%<br>90% |
| Genetics† (all participants, 160 cases and 840 controls per centre) | Risk locus | MAF 0.30<br>MAF 0.15 | OR≥1.8<br>OR≥2.0<br>OR≥1.8<br>OR≥2.0 | >99%<br>>99%<br>>99%<br>>99% | 84%<br>98%<br>44%<br>77% |
| Proteomics (25 cases and 25 controls / centre) | Levels will likely be several-fold higher in those with early disease [7] | Power to will depend on the fold-change, variance and (pre-)analytical factors. Similar work has been conducted with comparable sample sizes [8] |  |  |  |

MAF: minor allele frequency. †Multiplicative model. As reverse causation is of a lesser concern for the genomics analyses, those with established renal dysfunction (an additional 5-10% of the population despite exclusion of those with self-reported CKD) will also be included as cases to increase power. Includes correction for false discovery rate at accepted genome-wide thresholds; All calculations were done in Stata (v16).
<sup>#</sup>Data from Nicaragua suggest >10% of participants lose eGFR at >5mL/min/1.7m<sup>2</sup>/year (excluding those with pre-existing CKD) so there should be at least 100 'cases' at each site when using a dichotomous outcome.

The power is excellent for dichotomous exposures, and will be considerably greater (>99%) for continuous variable analyses. The genomic analyses are only powered to detect strong genetic effects, although many renal traits do exhibit these.(35, 36) **We are focusing on hypotheses which have the potential to explain the epidemics**, which involve CKDu prevalence at least 5-10 times that of ‘background’ rates in the same regions, and roughly 100 times the rates seen in young adults in Western countries. **For a risk factor to explain the extremely high prevalence of CKDu in these areas, it therefore must have a high prevalence (with at least 25% of the population exposed) and strong relative risks of CKDu**. For example, when Aristolochic Acid was linked to the epidemics of Balkan Nephropathy, it involved relative risks of 5-10 times(37, 38). Thus, we give a range of possible effect estimates, but **it is the power for the stronger odds ratios (i.e. OR=4) that is most relevant**.

## References

1. Chatterjee R. Occupational hazard. Science. 2016;352(6281):24–7.

2. Pearce N, Caplin B, Gunawardena N, Kaur P, O’Callaghan-Gordo C, Ruwanpathirana T. CKD of unknown cause: a global epidemic? Kidney International Reports. 2019;4:367–9.

3. Jha V, Garcia-Garcia G, Iseki K, Li Z, Naicker S, Plattner B, et al. Chronic kidney disease: global dimension and perspectives. Lancet. 2013;382(9888):260–72.

4. Gonzalez-Quiroz M, Pearce N, Caplin B, Nitsch D. What do epidemiological studies tell us about the chronic kidney disease of undetermined cause in Maso-America? A systematic review and meta-analysis. Chronic Kidney Journal. 2018;11:496–506.

5. Caplin B, Jakobsson K, Glaser J, Nitsch D, Jha V, Singh AK, et al. International Collaboration for the Epidemiology of eGFR in Low and Middle Income Populations – Rationale and core protocol for the Disadvantaged populations eGFR epidemiology study (DEGREE). BMC Nephrology 2016 18:1; DOI 10.1186/s12882-016-0417-1.

6. Yakub F. Kidney disease in farming communities remains a mystery. The Lancet. 2014;383:1794–5.

7. Kaur P, Gunawardena N, Kumaresan J. A review of chronic kidney disease of unknown etiology in Sri Lanka, 2001-2015. Indian Journal of Nephrology. 2019:in press.

8. Pearce N, Caplin B. Let’s take the heat out of the CKDu debate: more evidence is needed. Occupational and Environmental Medicine. 2019;76:357–9.

9. Rutter CE, Njoroge M, Cooper PJ, Prabhakaran D, Jha V, Kaur P, et al. International prevalence patterns of low eGFR in adults aged 18-60 without traditional risk factors from a population-based cross-sectional disadvantaged populations eGFR epidemiology (DEGREE) study. Kidney Int. 2025;107(3):541–57.

10. Correa-Rotter R, Wesseling C, Johnson RJ. CKD of unknown origin in Central America: the case for a Mesoamerican nephropathy. Am J Kidney Dis. 2014;63(3):506–20.

11. Wijkström J, González-Quiroz M, Hernandez M, Trujillo Z, Hultenby K, Ring A, et al. Renal Morphology, Clinical Findings, and Progression Rate in Mesoamerican Nephropathy. Am J Kidney Dis. 2017;69(5):626–36.

12. Herath C, Jayasumana C, De Silva P, De Silva PHC, Siribaddana S, De Broe ME. Kidney Diseases in Agricultural Communities: A Case Against Heat-Stress Nephropathy. Kidney International Reports. 2018;3(2):271–80.

13. Gallo-Ruiz L, Sennett CM, Sánchez-Delgado M, García-Urbina A, Gámez-Altamirano T, Basra K, et al. Prevalence and Risk Factors for CKD Among Brickmaking Workers in La Paz Centro, Nicaragua. Am J Kidney Dis. 2019;74(2):239–47.

14. Riefkohl A, Ramirez-Rubio O, Laws RL, McClean MD, Weiner DE, Kaufman JS, et al. Leptospira seropositivity as a risk factor for Mesoamerican Nephropathy. International Journal of Occupational and Environmental Health. 2017;23(1):1–10.

15. Torres C, Aragon A, Gonzalez M, Lopez I, Jakobsson K, Elinder CG, et al. Decreased Kidney Function of Unknown Cause in Nicaragua: A Community-Based Survey. American Journal of Kidney Diseases. 2010;55(3):485–96.

16. Gonzalez-Quiroz M, Smpokou E-T, Silverwood RJ, Camacaho A, Faber D, Garcia BLR, et al. Marked decline in kidney function amongst apparently healthy young adults at risk of Mesoamerican nephropathy. Journal of the American Society of Nephrology. 2018;29:2200–12.

17. Caplin B, Anand S, González-Quiroz M, Madero M, Michael M, Mohan S, et al. Taking the “unknown” out of CKDu-optimizing approaches to uncover the cause(s) of epidemic-level kidney disease in low- and middle-income settings: a report from the ISN’s International Consortium of CKDu Collaborators (ISN i3C). Kidney Int. 2026;109(4):652–60.

18. Wikstrom J. Morphological and clinical findings in Sri Lankan patients with chronic kidney disease of unknown cause (CKDu): Similarities and differences with Mesoamerican Nephropathy. PLoS Medicine. 2018.

19. Vervaet BA, Nast CC, Jayasumana C, Schreurs G, Roels F, Herath C, et al. Chronic interstitial nephritis in agricultural communities is a toxin-induced proximal tubular nephropathy. Kidney Int. 2020;97(2):350–69.

20. Wikstrom J. Kideny International. 2020:786–7.

21. Wimalawansa SJ. Renal tubular lysosomal vacuoles are a generic toxic manifestation and not particularly associated with agrochemicals and heavy metal toxicity or specific to a disease. Kidney Int. 2020;97(5):1058.

22. Badurdeen Z, Nanayakkara N, Ratnatunga NVI, Wazil AWM, Abeysekera TDJ, Rajakrishna PN, et al. Chronic kidney disease of uncertain etiology in Sri Lanka is a possible sequel of interstitial nephritis! Clinicopathological profile of symptomatic, newly-diagnosed CKDu patients. Clinical Nephrology. 2016;86:S106–S9.

23. Fischer RSB, Vangala C, Mandayam S, Chavarria D, Garcia-Trabanino R, Garcia F, et al. Clinical markers to predict progression from acute to chronic kidney disease in Mesoamerican nephropathy. Kidney International. 2018;94(6):1205–16.

24. Gonzalez-Quiroz M, Nitsch D, Hamilton S, O’Callaghan-Gordo C, Saran R, Glaser J, et al. Rationale and Population-based prospective cohort protocol for the Disadvantaged Populations at Risk of Decline in eGFR (CO-DEGREE). BMJ Open. 2019;9:e031169

25. Gonzalez-Quiroz M, Camacho A, Faber D, Aragon A, Wesseling C, Glaser J, et al. Rationale, description and baseline findings of a community-based prospective cohort study of kidney function amongst the young rural population of Northwestern Nicaragua. BMC Nephrology. 2017;18:16; DOI 0.1186/s12882-016-0422-4.

26. Ruwanpathirana T, Senanayake S, Gunawardena N, Munasinghe A, Ginige S, Gamage D, et al. Prevalence and risk factors for impaired kidney function in the district of Anaradhapura, Sri Lanka. BMC Public Health. 2019;19:763.

27. Checkoway H, Pearce N, Kriebel D. Research methods in occupational epidemiology. 2 ed. New York: Oxford University Press; 2004.

28. Soares AA, Eyff TF, Campani RB, Ritter L, Camargo JL, Silveiro SP. Glomerular filtration rate measurement and prediction equations. Clin Chem Lab Med. 2009;47(9):1023–32.

29. Smpokou E-T, Gonzalez M, Martins C, Le Blond J, Glaser J, Aragon A, et al. Systematic investigation of environmental exposures in young adults with declining kidney function in a population at risk of Mesoamerican Nephropathy. Occupational and Environmental Medicine. 2019:920–6.

30. Gorski M, Tin A, Garnaas M, McMahon GM, Chu AY, Tayo BO, et al. Genome-wide association study of kidney function decline in individuals of European descent. Kidney International. 2015;87(5):1017–29.

31. Yang J, Zaitlen NA, Goddard ME, Visscher PM, Price AL. Advantages and pitfalls in the application of mixed-model association methods. Nature Genetics. 2014;46(2):100–6.

32. Charlton JR, Portilla D, Okusa MD. A basic science view of acute kidney injury biomarkers. Nephrology Dialysis Transplantation. 2014;29(7):1301–11.

33. Gonzalez-Quiroz M, Smpokou E-T, Pearce N, Caplin D, Nitsch D. Identification of young adults at risk of an accelerated loss of kidney function in an area affected by Mesoamerican nephropathy BMC Nephrology. 2019;20:21.

34. Greenland S, Daniel R, Pearce N. Outcome modelling strategies in epidemiology: traditional methods and basic alternatives. International Journal of Epidemiology. 2016;45(2):565–75.

35. Gale DP, Molyneux K, Wimbury D, Higgins D, Levine AP, Caplin B, et al. Galactosylation of IgA1 Is Associated with Common Variation in C1GALT1. Journal of the American Society of Nephrology. 2017;28:2158–66.

36. Stanescu HC, Arcos-Burgos M, Medlar A, Bockenhauer D, Kottgen A, Dragomirescu L, et al. Risk HLA-DQA1 and PLA(2)R1 alleles in idiopathic membranous nephropathy. N Engl J Med. 2011;364(7):616–26.

37. Stiborova M, Arlt VM, Schmeiser HH. Balkan endemic nephropathy: an update on its aetiology. Archives of Toxicology. 2016;90(11):2595–615.

38. Tatu CA, Orem WH, Finkelman RB, Feder GL. The etiology of Balkan endemic nephropathy: Still more questions than answers. Environmental Health Perspectives. 1998;106(11):689–700.

